# Neoadjuvant and Adjuvant Pembrolizumab in Resectable Locally Advanced, Human Papillomavirus-Unrelated Head and Neck Cancer: A Multicenter, Phase 2 Trial

**DOI:** 10.1101/2020.03.18.20037846

**Authors:** Ravindra Uppaluri, Katie M. Campbell, Ann Marie Egloff, Paul Zolkind, Zachary L. Skidmore, Brian Nussenbaum, Randal C. Paniello, Jason T. Rich, Ryan Jackson, Patrik Pipkorn, Loren P. Michel, Jessica Ley, Peter Oppelt, Gavin P. Dunn, Erica K. Barnell, Nicholas C. Spies, Tianxiang Lin, Tiantian Li, David T. Mulder, Youstina Hanna, Iulia Cirlan, Trevor J. Pugh, Tenny Mudianto, Rachel Riley, Liye Zhou, Vickie Jo, Matthew Stachler, Glenn J. Hanna, Jason Kass, Robert Haddad, Jonathan D. Schoenfeld, Evisa Gjini, Ana Lako, Wade Thorstad, Hiram A. Gay, Mackenzie Daly, Scott J. Rodig, Ian S. Hagemann, Dorina Kallogjeri, Jay F. Piccirillo, Rebecca D. Chernock, Malachi Griffith, Obi L. Griffith, Douglas R. Adkins

## Abstract

**Background:** Pembrolizumab improved survival of patients with recurrent or metastatic head and neck squamous-cell carcinoma (HNSCC). The aims of this phase 2 trial were to determine if pembrolizumab administered to patients with resectable locally advanced, human papillomavirus (HPV)-unrelated HNSCC would be safe, result in pathologic tumor response (pTR), and lower the relapse rate.

**Methods:** Neoadjuvant pembrolizumab (200 mg) was administered 2-3 weeks before surgery. Resection of the primary tumor and involved/at-risk nodes was performed. Post-operative (chemo) radiation was planned. Patients with high-risk pathology (positive margins and/or extranodal extension) were to receive adjuvant pembrolizumab. pTR was quantified as the proportion of the resection bed with tumor necrosis, keratinous debris, and giant cells/histiocytes: pTR-0 (<10%), pTR-1 (10-49%), and pTR-2 (≥50%). Co-primary endpoints were pTR-2 among all patients, and one-year relapse rate in patients with high-risk pathology (historical: 35%). Correlations of baseline PD-L1 expression and T-cell infiltration with pTR were assessed, and tumor clonal dynamics were evaluated. Analyses were done per protocol. This trial is registered with ClinicalTrials.gov(NCT02296684), and is ongoing but closed to accrual.

**Findings:** Between June 30, 2015, and March 30, 2018, 36 patients enrolled. After neoadjuvant pembrolizumab, serious (grades 3-4) adverse events and unexpected surgical delays/complications did not occur. pTR-2 occurred in eight patients (22%), and pTR-1 in eight other patients (22%). pTR ≥10% correlated with baseline tumor PD-L1 expression, immune infiltrate, and IFN-γ pathway activity. Matched sample analysis showed compensatory upregulation of multiple immune inhibitory checkpoints in patients with pTR-0, and confirmed that clonal loss occurred in some patients. The one-year relapse rate among the eighteen patients with high-risk pathology was 16.7% (95%CI: 3.6-41.4%).

**Conclusions:** Among patients with locally advanced, HPV-unrelated HNSCC, neoadjuvant pembrolizumab was safe, and resulted in pTR-1 or pTR-2 in 44% of patients. The one-year relapse rate in patients with high-risk-pathology was lower than historical.

**Funding:** Merck, NCI, NIDCR, NHGRI and The V Foundation.

## INTRODUCTION

Patients with head and neck squamous-cell carcinoma (HNSCC) usually present with locally advanced disease. Surgery is the initial treatment in many of these patients. A large subset of surgically treated patients have high-risk pathology features (positive margin and/or extranodal extension) that is best treated with intensive post-operative adjuvant cisplatin and radiation therapy (POACRT). However, 35% of patients, particularly those with human papillomavirus (HPV)-unrelated HNSCC, will develop relapse of disease.^1,2^ Attempts to improve this outcome have been unsuccessful.^3^ Novel treatment strategies are needed for these patients.

Immune checkpoint inhibitors could reduce the risk of disease relapse in resectable locally advanced, HPV-negative HNSCC. Randomized trials showed that pembrolizumab and nivolumab, inhibitors of the programmed death receptor-1 (PD-1), improved overall survival (OS) of patients with platinum-resistant HNSCC.^4,5^ Pembrolizumab given alone or with chemotherapy improved the OS of patients with untreated recurrent or metastatic disease.^6^ The results of these trials provide strong rationale for evaluating the clinical impact of immune checkpoint inhibitors in patients with resectable locally advanced, HPV-unrelated HNSCC.

Immunotherapy may be administered before (neoadjuvant) or after (adjuvant) surgery. Pre-clinical experiments with several cancer types showed that administration of immunotherapy in the neoadjuvant interval provided greater benefit than when given in the adjuvant interval.^7–9^ In mouse models of spontaneously metastatic breast cancer, neoadjuvant immunotherapy and surgery were more effective in generating tumor-specific CD8+ T-cells and preventing development of lethal metastases than surgery alone.^7,8^ Importantly, the benefit of immunotherapy was dependent on resection of the primary tumor. In syngeneic mouse models of HPV-unrelated oral cavity carcinoma with defined T-cell antigens, administration of PD-1 inhibitor before, but not after, surgery reversed functional immunodominance, and induced effector T-cell immunity that resulted in rejection of tumor re-challenge after surgery.^9^ Collectively, these data support the critical importance of coordinating administration of immunotherapy before surgery to achieve optimal disease control with this novel strategy.

In addition to improved disease control, administration of immune checkpoint inhibitors in the neoadjuvant interval could result in other clinical benefits, including reduction of the rate of high-risk pathology and downstaging of the cancer. These outcomes could alter the selection of adjuvant therapy, resulting in less intense treatment. Finally, matched tumor tissue obtained at baseline and at surgery can be evaluated to define biomarkers that predict tumor response and resistance to immune checkpoint inhibitors. HNSCC is ideally suited for testing the effect of administration of immune checkpoint inhibitors in the neoadjuvant interval. Tumor is readily accessible to obtain matched baseline and surgical tissue, and to perform visual assessments of tumor response.^10^ Although reported in a few other cancers^11–17^, the safety and biologic and clinical effects of administration of immune checkpoint inhibitors in the neoadjuvant interval have not been reported in patients with resectable locally advanced HNSCC.

In this multicenter, phase 2 trial, we aimed to determine if administration of neoadjuvant pembrolizumab to patients with resectable locally advanced, HPV-unrelated HNSCC would be safe and result in pathologic tumor responses (pTR). We evaluated the immunologic correlates to pTR and assessed tumor clonal dynamics in matched tumor samples obtained before and after neoadjuvant pembrolizumab. Among patients with high-risk pathology, we aimed to determine if administration of neoadjuvant and adjuvant pembrolizumab would result in a relapse rate lower than historical. Herein, we report the results of our trial.

## METHODS

### Study Design and Participants

We did a multicenter, phase 2 trial at two university sites in the USA: Washington University, St. Louis, MO and Dana-Farber/Brigham and Women’s Cancer Center, Boston, MA. The trial is a non-randomized two-group study. Patient selection criteria were the same for both groups. Group 1 completed enrollment before Group 2 accrued patients. Group 1 is reported here. In group 1, patients were treated with one dose of neoadjuvant pembrolizumab, followed by surgery.

Patients with high-risk pathology were scheduled to be treated with POACRT followed by adjuvant pembrolizumab; patients without high-risk pathology were scheduled to be treated with post-operative adjuvant radiation therapy (POART) or observation but not adjuvant pembrolizumab. In group 2, patients were treated with two doses of neoadjuvant pembrolizumab, followed by surgery, then POACRT if high risk pathology or POART (or observation) if without high-risk pathology. Group 2 did not receive adjuvant pembrolizumab. Group 2 is ongoing, having accrued 25 of the planned sample size of 31 patients; the data are not yet mature. The results of group 2 will be reported at a later date.

Eligible patients had resectable clinical stage III-IVb (AJCC, 7^th^ Edition), HPV-unrelated (oral cavity, larynx, hypopharynx or p16-negative oropharynx) HNSCC, measurable disease per Response Evaluation Criteria in Solid Tumors (RECIST)v1.1, Eastern Cooperative Oncology Group (ECOG) performance status 0-1, and adequate marrow and organ function (absolute neutrophil count ≥1,500/mcL, platelets ≥100,000/mcL, hemoglobin ≥9g/dL; total bilirubin ≤1.5x upper limits of normal [ULN], AST and ALT ≤2.5x ULN; serum creatinine ≤1.5x ULN or creatinine clearance ≥30 mL/min). Key exclusion criteria included HPV-related oropharynx SCC, active autoimmune disease, or immunodeficiency.

Tests required to determine eligibility included complete blood count, metabolic panel, pregnancy test (women), coagulation and thyroid panels, urinalysis, and CT scans of the neck and chest.

The study protocol was approved by the institutional review board at each participating site. All patients provided written, signed, informed consent to participate. Independent data monitoring was done by the quality assurance committee of Washington University (St. Louis, MO, USA). The protocol is in the appendix (see Online).

### Procedures

In group 1, patients received one dose of pembrolizumab (200 mg, IV) 13-22 days before (neoadjuvant) surgery. Surgery included resection of all gross disease at the primary site, ipsilateral (and contralateral, in some patients) therapeutic/prophylactic neck dissection, and reconstruction using pedicled or free-flap procedures as deemed appropriate. Surgical tumor resection margins were defined by baseline assessments obtained before neoadjuvant pembrolizumab.

Patients with high-risk pathology were scheduled to be treated with POACRT, if they had adequate organ function and had recovered from surgery.^1,2^ Upon resolution of POACRT-related adverse events (AEs) to ≤ grade one and after three months from surgery date, patients with high-risk pathology were scheduled to be treated with adjuvant pembrolizumab (200 mg) every three weeks for six doses. Dose delays of pembrolizumab and treatment of immune-related AEs were performed per protocol. Patients without high-risk pathology (low/intermediate-risk) were scheduled to be treated with POART (or observation), but not adjuvant pembrolizumab. Physician discretion in selection of standard of care adjuvant therapy was permitted in patients with intermediate-risk pathology, such that some patients were treated with POACRT even though they lacked traditional high-risk-pathology. Administration of POA(C)RT was performed per protocol.

Before neoadjuvant pembrolizumab, patients underwent baseline assessment by physical examination and CT scan of the neck, and a biopsy of the primary tumor and collection of peripheral blood for correlative studies. Patients were assessed by physical examination the day before or day of surgery. After patient 20, the trial was amended to perform a CT scan of the neck within ten days prior to surgery. On the day of surgery, tissue from the primary tumor and peripheral blood were collected for correlative studies. AEs were assessed using revised Common Terminology Criteria for Adverse Events (CTCAE)v4.0. Peri-surgical AEs were assessed using Clavien-Dindo Classification of Surgical Complications.^18^ Tumor response was assessed using RECISTv1.1. Patients were monitored for 30 days after surgery for AEs and surgical/wound healing complications. During the administration of adjuvant pembrolizumab, patients were assessed every three weeks beginning with the first dose of pembrolizumab by physical examination, complete blood count, and metabolic panel, and every six weeks by thyroid panel and urinalysis. In these patients, AEs were monitored for 90 days after the last dose of adjuvant pembrolizumab. Patients were monitored for relapse every three months after surgery by physical examination and CT scans.

Details of PD-L1 immunohistochemistry (IHC) and multiplex immunofluorescence (MIF) are described in the appendix (see Online). Details regarding whole exome sequencing (WES), performed on tumor samples and matched normal blood, RNA sequencing (RNAseq) on tumor samples, T cell receptor sequencing (CapTCR-seq)^19^ on peripheral blood, multisector targeted genome sequencing on tumor samples, and analyses (expression, deconvolution, TCR repertoire, etc.) are provided in the **Supplementary Appendix**. DNA and RNA data were processed using the Genome Modeling System.^20,21^ Sequencing data have been deposited in dbGaP (phs001623). Patient HLA haplotypes and mutational profiles were used to predict putative neoantigens with pVACtools.^22^

#### Outcomes

The co-primary endpoints were one-year relapse rate (the absolute proportion of patients who developed local-regional and/or distant relapse within one year of surgery) in patients with high-risk pathology, and the proportion of all patients with pTR-2 in the surgical specimen after administration of one dose of neoadjuvant pembrolizumab. pTR was defined as the presence of tumor cell necrosis and keratinous debris with giant cell/histiocytic reaction, quantified as a percentage of the overall tumor bed (area pathologic response/area pathologic response plus viable tumor): pTR-0 (<10%), pTR-1 (10-49%), and pTR-2 (≥50%). Two pathologists with head and neck expertise (RDC and IH) independently evaluated all slides from baseline and surgery specimens and quantified pTR in increments of 10%. Primary tumor and lymph node metastases were scored separately. Overall pTR was classified based on the best pTR observed in either primary tumor or lymph node. Joint review consensus was reached when discrepancies occurred. Current standardized definitions of immune-checkpoint inhibitor-induced pTR were not available when the study was begun, and were not used in the analysis. Secondary endpoints included safety of administration of neoadjuvant pembrolizumab, and clinical tumor response to neoadjuvant pembrolizumab assessed by physical examination and, in some patients, by RECISTv1.1. Correlative endpoints assessed on matched tumor specimens obtained before and after (on day of surgery) neoadjuvant pembrolizumab included PD-L1 expression, histologic, immunologic, genomic, and tumor clonal dynamic changes. T-cell clonality was performed on peripheral blood obtained before and after neoadjuvant pembrolizumab (**Supplementary Figure 1, Supplementary Table 1**).

**Table 1.** Patient Characteristics by Pathological Risk Category.

| Characteristic | All Patients (N=36) | Patients with High-Risk Pathology (N=18) | Patients with Low/Int.-Risk Pathology (N=18) | p-value* | Diff (95% CI) |
| --- | --- | --- | --- | --- | --- |
| **Age at enrollment, y** |  |  |  |  |  |
| Median (range) | 60 (32-87) | 61.5 (37-87) | 58.5 (32-73) | 0.16 | 5 (-2 to 13) |
| **Sex, N (%)** |  |  |  |  |  |
| Male | 26 (72) | 13 (72) | 13 (72) | 1.00 | 0 (-29 to 29) |
| Female | 10 (28) | 5 (28) | 5 (28) |  |  |
| **Ethnicity** |  |  |  |  |  |
| White | 28 (78) | 15 (83) | 13 (72) | 0.82 | 11 (-16 to 38) |
| Black | 6 (17) | 2 (11) | 4 (22) |  | -11 (-35 to 13) |
| Asian | 2 (5) | 1 (6) | 1 (6) |  | 0 (-15 to 15) |
| **Smoking history, N (%)** |  |  |  |  |  |
| Ever | 31 (86) | 16 (89) | 15 (83) | 1.00 | 6 (-17 to 28) |
| Never | 5 (14) | 2 (11) | 3 (17) |  |  |
| **Smoking pack-years** |  |  |  |  |  |
| Median (range) | 30 (0-80) | 30 (0-80) | 30 (0-80) | 0.79 | 0 (-15 to 10) |
| **Alcohol use history** |  |  |  |  |  |
| Ever | 23 (64) | 11 (61) | 12 (67) | 1.00 | -6 (-37 to 26) |
| Never | 13 (36) | 7 (39) | 6 (33) |  |  |
| **ECOG performance status, N (%)** |  |  |  |  |  |
| 0 | 26 (72) | 16 (89) | 10 (56) | 0.06 | 33 (6 to 60) |
| 1 | 10 (28) | 2 (11) | 8 (44) |  |  |
| **Tumor site** |  |  |  |  |  |
| Larynx/Hypopharynx | 10 (28) | 4 (22) | 6 (33) | 0.56 | -11 (-4 to 18) |
| Oral Cavity | 22 (61) | 11 (61) | 11 (61) |  | 0 (-32 to 32) |
| Oropharynx | 4 (11) | 3 (17) | 1 (6) |  | 11 (-9 to 31) |
| **Clinical T stage, N (%)** |  |  |  |  |  |
| T1-T2 | 7 (19) | 4 (22) | 3 (17) | 0.59 | 5 (-20 to 31) |
| T3 | 4 (11) | 3 (17) | 1 (6) |  | 11 (-9 to 31) |
| T4 | 25 (69) | 11 (61) | 14 (78) |  | -17 (-46 to 13) |
| **Clinical N stage, N (%)** |  |  |  |  |  |
| N0-N1 | 13 (36) | 2 (11) | 11 (61) | 0.005 | -50 (-77 to 23) |
| N2 | 21 (58) | 14 (78) | 7 (39) |  | 39 (9 to 68) |
| N3 | 2 (6) | 2 (11) | 0 |  | 11 (-3 to 26) |
| **Clinical disease stage, N (%)** |  |  |  |  |  |
| III | 3 (8) | 0 | 3 (17) | 0.23 | -17 (-34 to 1) |
| IV | 33 (92) | 18 (100) | 15 (83) |  |  |
| **Days first visit to surgery** |  |  |  |  |  |
| Median (range) | 33 (25-76) | 35 (25-76) | 33 (25-58) | 0.81 | 1 (-5 to 11) |
| **Days neoadjuvant pembrolizumab to surgery** |  |  |  |  |  |
| Median (range) | 16 (13-22) | 16 (13-22) | 18 (14-22) | 0.23 | -1 (-3 to 1) |
| **Pathologic disease stage, N (%)** |  |  |  |  |  |
| I-II | 3 (8) | 0 | 3 (17) | 1.00 | -17 (-34 to 1) |
| III | 6 (17) | 2 (11) | 4 (22) |  | -11 (-35 to 13) |
| IVA-IVB | 27 (75) | 16 (89) | 11 (61) |  | 28 (10 to 55) |

\*Wilcoxon signed rank test used for continuous variables; Fisher's exact test used for categorical variables

**Figure 1.**
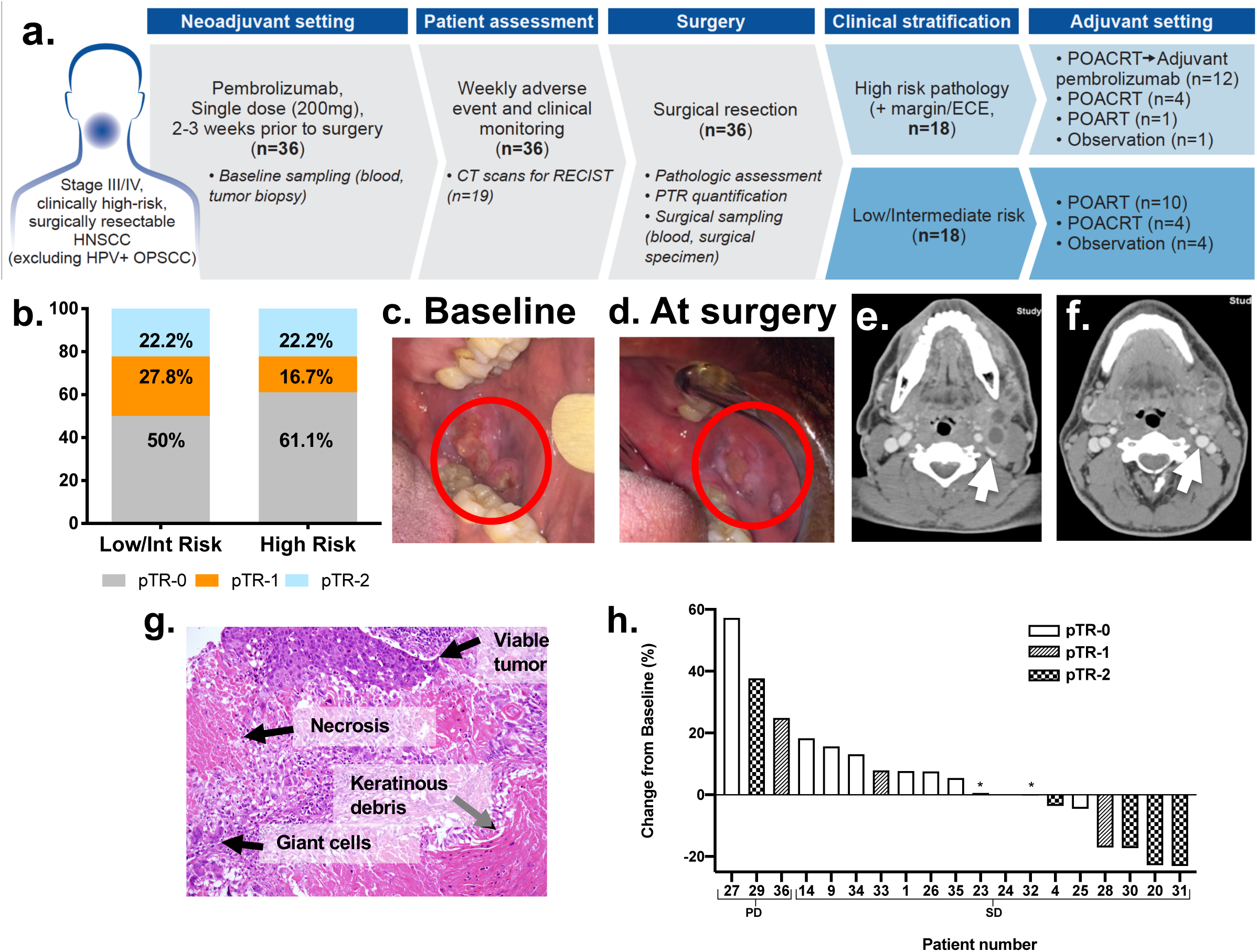
Trial profile and tumor responses (pathologic and radiologic) to neoadjuvant pembrolizumab. (A) Trial profile: Patients (n=36) with locally advanced, Stage III/IV, HPV-negative HNSCCs underwent baseline tumor and blood sampling and received neoadjuvant pembrolizumab 2-3 weeks before surgery. Of the 18 patients with high-risk pathology, 12 received adjuvant pembrolizumab. Patients with low/intermediate-risk pathology did not receive adjuvant pembrolizumab. (B) pathologic tumor response-2 (pTR-2) was observed in similar proportions of patients with low/intermediate and high-risk pathology. (C) Baseline and (D) post-neoadjuvant treatment (at surgery) images of oral cavity primary cancer in Patient 20 showing dramatic decrease in the size. (E) Representative CT images at baseline and (F) after neoadjuvant pembrolizumab (day prior to surgery), confirmed tumor response seen on physical exam. Notably, the pre-treatment CT and FDG-PET/CT scan (not shown) showed multiple large and necrotic FDG-avid neck lymph nodes, which are radiologic signs of SCC. Of note, the internal jugular vein (white arrow) was compressed on the baseline scan (E) and appeared fuller on the post-treatment scan (F). (G) Representative H&E slide of pTR highlighting changes noted in surgical specimens. (H) Nineteen patients had CT evaluations at baseline and prior to surgery following neoadjuvant pembrolizumab: 16 with stable disease (SD) and 3 with progressive disease (PD) by RECIST criteria. *Indicates two patients with pTR-1.

No major protocol deviations occurred. The protocol was amended six times over the course of the study. Amendments 1-3 included updates of the risk profile and dose modifications of study drug, clarified eligibility criteria, and added Dana-Farber/Brigham and Women’s Cancer Center, Boston, MA as a secondary site. Amendment 4 added correlative studies. Amendment 5 added an unplanned interim analysis after the first 20 patients enrolled into group 1 due to the lower-than-expected rate of patients with high-risk pathology, and added CT scan of the neck to be performed after neoadjuvant pembrolizumab and prior to surgery. Amendment 6 closed accrual to group 1, and added group 2.

#### Statistical Analysis

Relapse rate at one year in patients with high-risk pathology was the initial primary endpoint. Historical data showed that the one-year relapse rate after surgery and POACRT in patients with high-risk pathology was 35%.^1,2^ Relapse rate at one year was selected because the majority (>90%) of relapse events in these patients occurred within one year of surgery.^1,2^ A sample size of 31 evaluable patients was required to detect a reduction in the one-year relapse rate to ≤20%, with a power of 80%, using a one-sided alpha of 0.05. Evaluable patients for this endpoint were those who had high-risk pathology in the surgical specimen after one dose of neoadjuvant pembrolizumab. Assuming a rate of high-risk pathology of 80% in patients with clinical stage III/IV HPV-unrelated HNSCC, and a 20% drop-out rate, we planned to accrue a total of 46 patients to group 1. However, after enrollment of the first 20 patients, the proportion of patients with high-risk pathology after one dose of neoadjuvant pembrolizumab and surgery was lower than expected (35%), prompting an unplanned interim analysis to assess the feasibility to achieve the initial primary endpoint. Enrollment continued during the interim analysis. The results of the interim analysis confirmed the unexpectedly lower rate of high-risk pathology. The trial was amended to: a) close enrollment of group 1 based on inability to accrue the required number of patients with high-risk pathology in a practical interval, b) addition of pTR-2 after neoadjuvant pembrolizumab in all patients as a co-primary endpoint, and c) addition of group 2, as previously described. In this report, the analysis of one-year relapse rate in patients with high-risk pathology was performed as intention-to-treat.

Stopping rules were in place for delay of surgery or serious (grade 3-5) AEs attributed to pembrolizumab. The study was to be stopped, and amended or closed, in the event of: 1) neoadjuvant pembrolizumab-related AEs leading to significant delay in surgery (more than 14 days delay in 1 of the first 15 patients, 2 of 30, or 3 of 45) or 2) serious pembrolizumab-related AEs (occurring in 1 of the first 10 patients, 2 of 20, 3 of 30, 4 of 40, or 5 of all patients).

Distribution of demographic and clinical characteristics was defined and compared between patients in high-risk and other (low/intermediate-risk) pathology groups. Percent difference and 95% confidence intervals (CI) were calculated for categorical variables; median difference and 95% CI were calculated for continuous variables. Spearman’s rank correlation coefficient was used to estimate correlations between tumor PD-L1 staining and numbers of tumor infiltrating T cells (CD8+ and CD4+) and extent of pTR. Molecular correlates were evaluated for changes across pTR categories using the non-parametric test of trend. Kaplan-Meier estimates of OS, progression-free survival (PFS) and relapse-free survival (RFS) rates and 95% CI by pathology risk category or pTR category were determined and differences between categories assessed using the log rank test. OS was defined as time (months) from day of surgery to death; PFS was defined as time from day of surgery to first disease progression event (new primary, recurrence, distant metastasis or death from disease) or death from any cause; RFS was defined as time from day of surgery to first relapse event (recurrence or distant metastasis). In gene expression analysis, unpaired Mann-Whitney/Wilcoxon rank-sum tests were used to compare groups of patients, and paired Wilcoxon signed-rank tests were used to compare matched baseline and post-treatment tumor samples.

The trial was registered with ClinicalTrials.gov (NCT02296684).

## RESULTS

Between June 30, 2015, and March 30, 2018, 36 patients enrolled into group 1 of the trial. Patient and tumor characteristics were typical of those observed in patients with locally advanced, HPV-unrelated HNSCC: mostly males with a smoking history, and large tumors (**Table 1**). The trial profile is shown in **Figure 1A**. All patients received one dose of neoadjuvant pembrolizumab and underwent surgery. Microvascular flap reconstruction was required in 28 patients. Eighteen patients (50%) had high-risk pathology, of which 12 were treated with POACRT and adjuvant pembrolizumab, 4 with POACRT, 1 with POART, and one was observed (**Supplementary Table 2**). Adjuvant pembrolizumab was not administered to 6 patients with high-risk pathology due to persistent toxicity of POACRT (2), patient decision (2), peri-operative myocardial infarction (1) and interim development of distant metastasis (1). Eighteen patients (50%) had low/intermediate-risk pathology, of which 10 were treated with POART, 4 with POACRT, and 4 were observed.

**Table 2.** Neoadjuvant and adjuvant pembrolizumab-related adverse events.

|  | **Neoadjuvant Phase** |  | **Post-Surgery and Adjuvant Phase** |  |
| --- | --- | --- | --- | --- |
|  | **All Patients**<br>**(N=36)** |  | **Patients Treated with Adjuvant**<br>**Pembrolizumab (N=12)\*** |  |
| **Event** | **Grades 1-2** | **Grades 3-4** | **Grades 1-2** | **Grades 3-4** |
|  | **N (%)** | **N (%)** | **N (%)** | **N (%)** |
| Fatigue | 2 (6) | 0 (0) | 2 (17) | 0 (0) |
| Hypothyroidism | 0 (0) | 0 (0) | 3 (25) | 1 (8) |
| Fever | 1 (3) | 0 (0) | 0 (0) | 0 (0) |
| Tumor flare | 1 (3) | 0 (0) | 0 (0) | 0 (0) |
| AST increase | 0 (0) | 0 (0) | 1 (8) | 0 (0) |
| ALT increase | 0 (0) | 0 (0) | 1 (8) | 0 (0) |
| Alk. Phos. increase | 0 (0) | 0 (0) | 1 (8) | 0 (0) |
| Diarrhea | 0 (0) | 0 (0) | 1 (8) | 0 (0) |

\*No post-surgery/adjuvant phase pembrolizumab-related adverse events occurred among patients who did not receive adjuvant pembrolizumab.

Administration of neoadjuvant pembrolizumab before surgery was safe. Serious immune-related AEs did not occur in the interval between administration of neoadjuvant pembrolizumab and through 30 days after surgery (**Table 2**). Unexpected surgical delays or complications were not observed (**Supplementary Table 3**). Delivery of POACRT was not compromised by prior neoadjuvant pembrolizumab. During administration of adjuvant pembrolizumab, one serious reversible immune-related AE occurred, hypothyroidism (**Table 2**).

Median follow-up after surgery was 22 months (IQR: 17.1-32.2); 97% had ≥ one year follow-up after surgery. In patients with high-risk pathology, the one-year relapse rate was 16.7% (3/18, 95%CI: 3.6-41.4). In patients with low/intermediate-risk pathology, the one-year relapse rate was 0% (**Supplementary Figures 2-3**).

**Figure 2.**
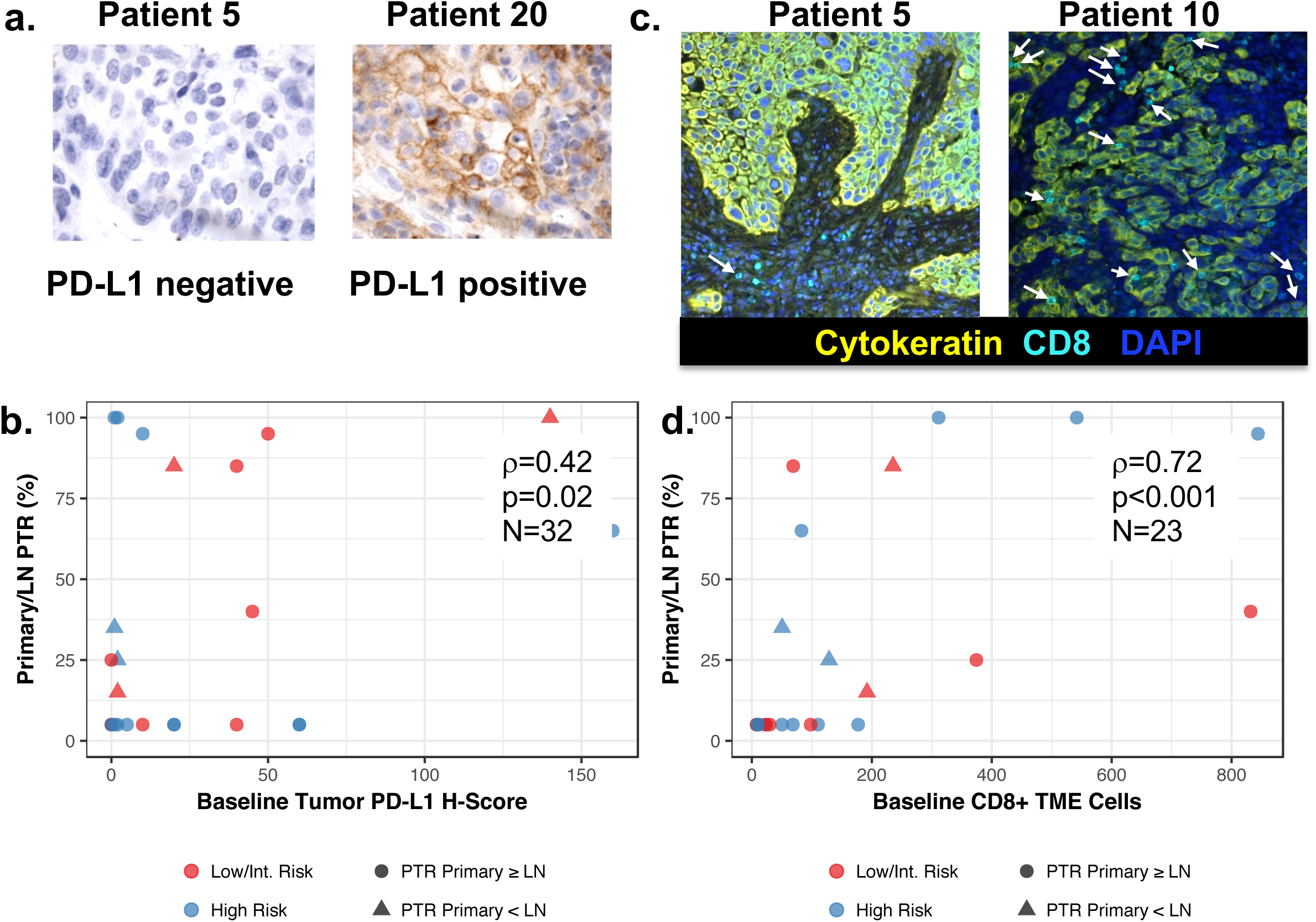
Pathologic tumor response correlates with tumor PD-L1 and immune infiltrates. (A) Representative PD-L1 staining of tumor biopsies at baseline. (B) PD-L1 H-score correlated with pathologic tumor response (PTR). Baseline PD-L1 primary tumor expression levels by IHC and percent PTR were significantly positively correlated for 32 evaluated patients (rho=0.43; 95%CI 0.079-0.668) (C) representative multiplex immunofluorescence (MIF) images showing patient 5 with minimal CD8+ T cell infiltrates and Patient 10 with higher CD8+ T cell infiltrates (white arrows) in baseline biopsies. (D) Extent of PTR was correlated with number of CD8+ T cells in the baseline biopsy tumor microenvironment (TME). Baseline number of TME CD8+ T cells assessed by MIF and percent PTR were significantly positively correlated for 23 evaluated patients (rho=0.72; 95%CI: 0.443-0.875).

After neoadjuvant pembrolizumab, pTR-2 occurred in the surgical specimens of 8 patients (22%) and pTR-1 occurred in 8 additional patients (22%). Overall, pTR of ≥10% was observed in 16 of 36 patients (44%) (**Figure 1B, Supplementary Table 4**). The proportion of patients who experienced pTR was similar in the high-risk and low/intermediate-risk pathology groups (**Figure 1B, Supplementary Table 5**). Of the patients with pTR and tumor in the primary site and lymph nodes, 8 of 10 had pTR in only one of these sites.

Significant clinical tumor responses occurred in a minor proportion of patients after neoadjuvant pembrolizumab. Patient 20 experienced an exceptional clinical tumor response, and tumor downstaging (clinical stage IV:T_2_N_2b_ downstaged to pathologic stage I:T_1_N_0_), after neoadjuvant pembrolizumab (**Figure 1C-F**). The surgical specimen from this patient showed extensive pTR (90%), and only a small residual focus of SCC (**Figure 1G**). Most patients had stable disease (**Figure 1H**). The proportion of patients with high-risk pathology (50%) was lower than expected (80%). Down-staging of cancer (defined as pathologic stage lower than clinical stage) after neoadjuvant pembrolizumab occurred in 7 patients (19%) (**Supplementary Table 4**).

We explored immunologic correlates of tumor response to neoadjuvant pembrolizumab using IHC, MIF, and RNAseq performed on baseline and surgical samples (**Supplementary Figure 1**). A strong positive correlation existed between PD-L1 protein expression in baseline biopsies and PTR (**Figures 2A-B**, p=0.02). MIF showed a strong positive correlation between extent of PTR and infiltration of CD8+ (r=0.72; 95%CI: 0.44-0.88) but not CD4+ T-cells (r=0.25; 95%CI: −0.16-0.62) in baseline biopsies (**Figures 2C-D, Supplementary Figure 4**). These immune cell populations were queried by RNAseq^17^ and revealed significantly higher levels of overall immune infiltrate, M1 macrophages, and CD4 and CD8 T cells (p<0.05) at baseline in patients with PTR compared to those without (**Figure 3, Supplementary Table 7**). Consistent with these findings, differential gene expression analysis revealed that patients with PTR (n=6) displayed significantly increased baseline expression of immune and inflammatory genes (e.g. *IFNG, CXCL9, CXCL10, CXCL11*, p<0.01, FDR<0.2) as well as significant enrichment of genes associated with these processes (p<0.01, FDR<0.2, **Figure 3A, Supplementary Table 8**), compared to patients without PTR (n=10). These inflammatory expression patterns were maintained in patients with PTR over treatment (n=4).

**Figure 3.**
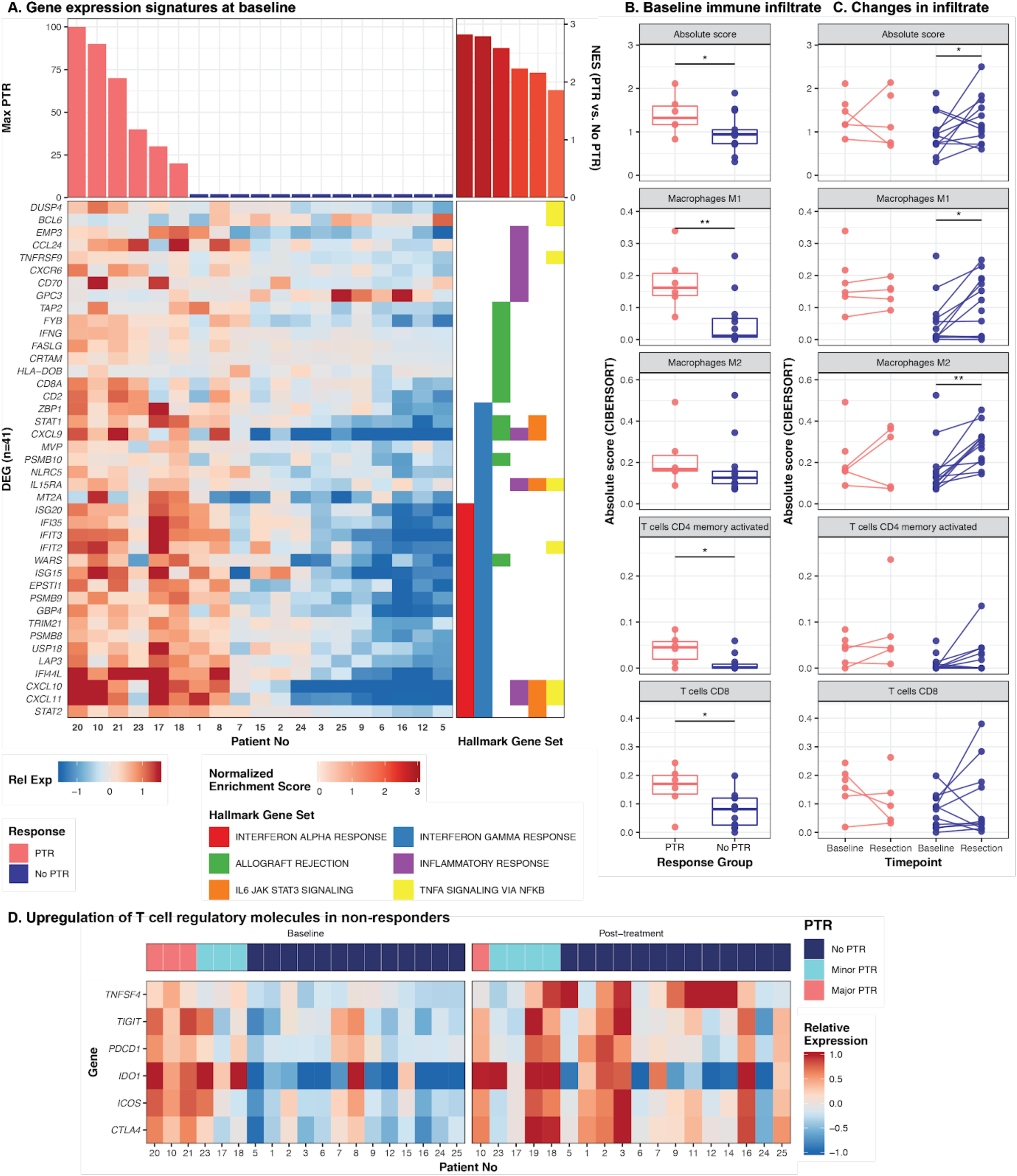
Immune infiltrate and activity correspond to patient response. **A**. Heatmap shows genes (n=41) associated with hallmark gene sets (right panel) that were differentially expressed (p<0.01, adjusted p<0.25) between patients with PTR and those without at baseline. Genes are sorted first by hallmark geneset, then by Ward’s hierarchical clustering. Patients are sorted by decreasing maximum PTR (at either the tumor or lymph node site), then by Ward’s hierarchical clustering. Expression is displayed as the gene-normalized expression across samples. **B-C**. Baseline- and post-treatment RNA was assessed for patterns of infiltrating immune cells, and values were summarized by the absolute levels of immune cells. These are indicated as the sum of all immune cell populations (Absolute score) or by subpopulation at baseline (B). Matched samples were available for a subset of patients (n=15) and the changes over the course of treatment are depicted by connected lines between baseline and resection timepoints in (C). Wilcoxon tests were used to evaluate statistical significance across responder groups and timepoints; * indicates p<0.05. **D**. Comparison of six genes (y-axis) that were upregulated post-treatment (right panel) in patients without PTR, compared to baseline (un-treated) samples (left panel). The heatmaps display the relative expression of genes (z-score; color). Patients were sorted by extent of PTR, then by unsupervised hierarchical clustering (with Ward’s ranked agglomeration).

**Figure 4.**
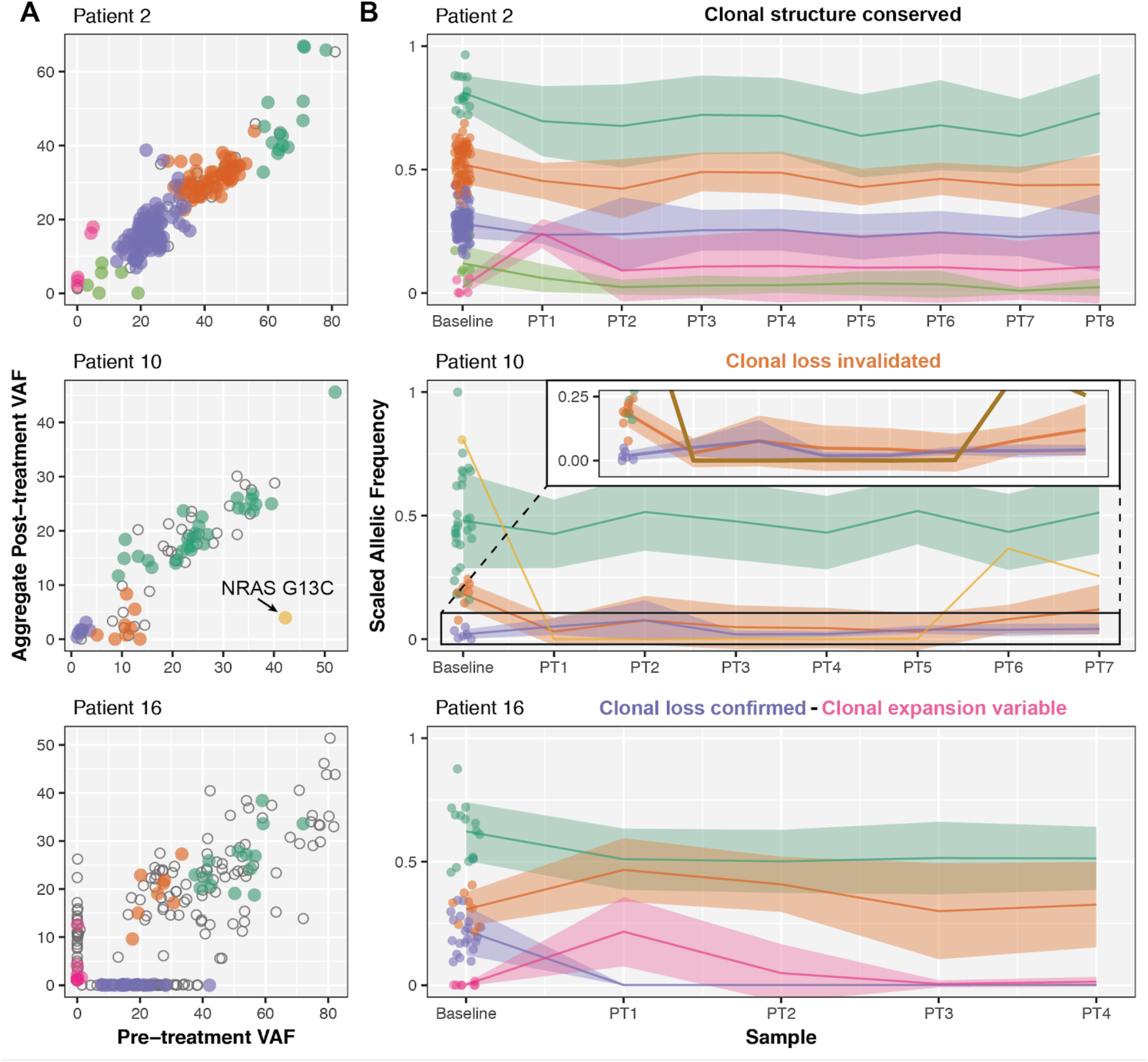
Varied clonal dynamics due to spatial heterogeneity. **A)** Clonality plots comparing VAF of SNVs and Indels at baseline (x-axis) and resection (y-axis). These values represent aggregate metrics of all post-treatment samples available, and colors represent variants that clustered together by comparing these aggregate metrics per individual. Open circles represent variants that were either unassigned or were assigned to clusters with less than 5 variants. **B)** The scaled allelic frequency (AF) is depicted on the y-axis of each variant (point) that was assigned a cluster in (A). The solid line represents the average scaled AF of the cluster. Shaded ribbons represent the standard error from the mean scaled AF of the cluster. Eight non-overlapping regions were isolated from Patient 2 post-treatment (PT) resected tumor, 7 regions were isolated from patient 10, and 8 regions were isolated from Patient 2 and analyzed (**Supplementary Figure 12**).

Interestingly, surgical samples from patients without PTR (n=11) after neoadjuvant pembrolizumab also showed enrichment for inflammatory gene signatures (p<0.01, FDR<0.2) which was counterbalanced by increased expression of T cell checkpoint molecules, including *PDCD1, CTLA4, ICOS, TIGIT, IDO1*, and *TNFSF4* (p<0.01, FDR<0.2, n=10, **Figure 3D, Supplementary Figure 5, Supplementary Table 8**).

TCR sequencing was performed to assess the peripheral blood T cell repertoire relationship to tumor-infiltrating immune-related expression patterns. There were no significant differences at baseline in the diversity, clonality, and richness in *TRB* clones of peripheral blood between patients without or with PTR. However, patients with PTR exhibited patterns associated with increased TCR diversity and clonality in peripheral blood after neoadjuvant pembrolizumab, with significantly higher Shannon, Gini Simpson, and Inverse Simpson diversity indices and lower geometric coefficient of variance (p<0.05, **Supplementary Table 9, Supplementary Figures 6-7**).^18^ We interrogated whether peripheral T cell clones may be tumor-associated, using bulk tumor RNA data (**Supplementary Appendix**), and identified 1,166 *TRA* clones (8-210 per patient) and 1,482 *TRB* clones (2-273 per patient) in tumor RNA. Of these, 62 *TRA* clones (0-21 per patient) and 88 *TRB* clones (0-39 per patient) were also detected in the peripheral T cell repertoire. Furthermore, these T cell clones detected in the bulk tumor tended to be of higher frequency in the peripheral blood, on average, ranking in the top 30.6% of clones detected in the peripheral blood (**Supplementary Figure 8**). Tumor-associated *TRB* clones were even more prevalent post-treatment, ranking in the top 40.0% of clones in baseline blood and the top 24.4% of clones in post-treatment peripheral blood.

We completed whole exome sequencing (WES) of DNA for 24 baseline and 22 surgical samples (25 patients, 21 matched pairs, average 91.3X coverage). Baseline tumor biopsies had nonsynonymous tumor mutation burden (TMB) ranging from 0.63-5.18 mutations/Mb (30-280 mutations), and the corresponding mutational landscape was consistent with that previously reported for HPV-unrelated HNSCC (**Supplementary Figure 9, Supplementary Table 10**).^19^ Neoantigens were predicted for each patient based upon their mutations^20^ and inferred HLA haplotypes^21^ (see **Methods** and **Supplementary Tables 11-12**). Baseline, neoantigen burden ranged from 0-163 (median 76) mutations. Baseline TMB and predicted neoantigen burden did not correlate with extent of PTR (r=-0.27, p=0.2 and r=-0.23, p=0.29, respectively; **Supplementary Figure 10**).

Comparing matched baseline and post-treatment tumor DNA, there were 2,431 variants (9-260 per patient, n=22 patients with matched timepoints) that were detected in both baseline and post-treatment samples, and 873 variants (2-116 per patient) that were detected uniquely at a single timepoint. Of these, 572 variants (1-101 per patient) were detected specifically in baseline tumor samples, and 301 variants (1-61 per patient) were detected specifically in post-treatment tumor samples. Variants detected solely at baseline could potentially indicate the event of immune rejection and clonal loss, while the emergence of variants post-treatment could indicate clonal expansion of a non-responsive tumor clone or the presence of *de novo* tumor subclones. However, we hypothesized that some of these clonal dynamics may be observed due to insufficient sequencing depth or sampling bias. To address the impact of sequencing depth on the observed clonal dynamics, we designed a targeted capture probeset on WES-defined mutations (mean 160X coverage) to achieve higher sequencing depth (mean 1,060X coverage; **Supplementary Tables 13-14**). Overall, variant allele fractions (VAFs) between WES and targeted capture sequencing were highly correlated (r=0.94). Higher depth sequencing was able to recover 198 variants that were originally found at only one timepoint by WES. However, there were still 675 variants (1-96 per patient) detected uniquely pre- or post-treatment, supporting the conclusion that neoadjuvant pembrolizumab may have induced clonal changes (**Supplementary Figure 11**).

Alternatively, if the baseline and post-treatment surgical sections came from different regions of the tumor, spatial heterogeneity could also explain the presence of variants at either timepoint. While additional baseline samples were unavailable, we performed multisector analysis by micro-dissecting surgical resection samples. FFPE-preserved tumor sections from 8 patients (41 samples, 3-8 per patient, **Supplementary Figure 12**) obtained from geographically distinct regions in all remaining tumor blocks were subjected to high-depth DNA sequencing using the custom capture probeset. Four patterns of tumor clonal dynamics were observed: (1) clonal conservation, (2) confirmation of clonal loss, (3) invalidation of clonal loss, and (4) observations of clonal expansion (in some or all regions surveyed).

In some patients, the mutational profile was strikingly maintained across all regions of the baseline and surgical resection (e.g. Patient 2, **Figure 4**), confirming the geographical conservation of clonal structure. In order to confirm the event of clonal loss, pre-treatment-specific variants that were originally defined would have to be completely absent across all regions of the post-treatment resected surgical specimen. There were 2-71 (median 6.5) variants completely undetected in the original post-treatment specimens (denoted ‘PT1’) across the 8 patients with multi-sector sequencing. This pattern was confirmed for 1-34 (median 3) variants across all available matched post-treatment specimens. Clonal loss was especially evident in Patients 15 (yellow cluster, **Supplementary Figure 13**) and 16 (purple cluster, **Figure 4**). On the other hand, there were 2-37 (median 6) variants recovered in at least one post-treatment sample across all 8 patients, invalidating some of our original observations of clonal loss. For example, in Patient 10 (orange and gold clusters, **Figure 4**), an *NRAS* G13C activating mutation (42.4% baseline DNA VAF) was recovered in 2/6 additional samples (PT6-7).

In the original procured samples, there were 0-25 (median 5.5) variants that were absent at baseline, suggesting the expansion of a novel or previously undetected subclone in 7 patients. Clonal expansion patterns were validated in Patients 1 and 15 (pink and gold cluster, respectively, **Supplementary Figure 13**), where some PT1-specific variants were detected across all post-treatment specimens. However, there were 4-49 variants (median 33) detected in only a subset of post-treatment samples. These patterns suggest that observations related to novel subclones or expansion are variable, due to spatial heterogeneity and sampling bias. For example, if PT3 or PT4 had originally been chosen for WES from Patient 16, the clonal expansion observed in PT1 would have been missed (see pink cluster, **Figure 4**). Furthermore, since there was spatial heterogeneity post-treatment, there is strong evidence that this heterogeneity also existed at baseline, but cannot be confirmed without further baseline sampling. Therefore we can not discount the possibility that apparent clonal expansions might be invalidated if spatial heterogeneity at baseline could be assessed.

## DISCUSSION

To the best of our knowledge, we report the first application of an immune checkpoint inhibitor as a therapeutic strategy in patients with locally advanced, HPV-unrelated head and neck cancer. Administration of neoadjuvant pembrolizumab was safe, and did not adversely affect the outcomes of surgery or the delivery of POACRT. Adjuvant pembrolizumab administered to patients with high-risk pathology was associated with a low risk of serious pembrolizumab-related AEs (8%).

In patients with locally advanced, HPV-unrelated HNSCC with high-risk pathology, we hypothesized that pembrolizumab administered before and after surgery would lower the rate of relapse. In our trial, the one-year relapse rate in the 18 patients with high-risk-pathology was 16·7%, lower than the historical rate of 35%.^1,2^ These results should be cautiously interpreted because the sample size was small and the upper limit of the 95%CI for one-year relapse rate included the historical relapse rate. An ongoing international phase 3 trial is testing the potential benefit of neoadjuvant and adjuvant pembrolizumab in patients with resectable, locally advanced HNSCC (NCT03765918).

In our trial, pTR-1 and pTR-2 occurred in 44% of patients after a single dose of neoadjuvant pembrolizumab. Patients with pTR ≥10% had better RFS compared to those without pTR. Although the sample size was small, these data provide the first evidence in patients with locally advanced HNSCC that pTR in response to neoadjuvant pembrolizumab may be a biomarker predictive of a lower rate of disease relapse. This hypothesis is being tested in an ongoing, phase 3 trial (NCT03765918). Pathologic complete response (pCR: absence of tumor in the surgical specimen) after one dose of neoadjuvant pembrolizumab did not occur. pCR rates of 15-45% were reported in other cancers^11–14^; however, these trials administered multiple doses of one or combinations of immune checkpoint inhibitors.

We report the most comprehensive evaluation of tumor clonal loss following neoadjuvant immunotherapy, based upon high-depth, multisector genome sequencing of surgical specimens. Depth of genome sequencing and spatial heterogeneity influenced interpretation of clonal dynamics. Using targeted genomic sequencing (mean 1,060X coverage) performed in multiple sectors of the surgical specimen, we confirmed that tumor clonal loss occurred after neoadjuvant pembrolizumab in some cases. In other cases, while WES of a single area showed tumor clonal loss, targeted genome sequencing of multiple sectors showed persistence of these tumor clones. Documentation of tumor clonal loss by high-depth, multisector genome sequencing in some patients is further evidence of the anti-tumor effect of neoadjuvant pembrolizumab.

Patients with pTR after neoadjuvant pembrolizumab had pre-existing primed immune effectors that were maintained over the course of treatment. pTR after neoadjuvant pembrolizumab correlated with baseline tumor PD-L1 expression, immune infiltrate, and IFN-γ pathway activity, but not with TMB. The lack of association of TMB and tumor response in our cohort may be due to limited patient numbers. In other cancers, tumor response to anti-PD-1 therapy correlated with TMB and with metrics which described the baseline T cell repertoire, infiltrating immune cell subpopulations and expression patterns related to their activity.^8,24–27^

Our study is the first to provide insights into mechanisms of intrinsic resistance to anti-PD1 therapy in patients with locally advanced HNSCC. Patients who did not experience pTR after neoadjuvant pembrolizumab showed limited immune activity in the baseline tumor specimen. A subset of these patients showed upregulation of immune pathways in the surgical sample after neoadjuvant pembrolizumab; however, this was counterbalanced by increased expression of T cell checkpoint molecules, including *PDCD1, CTLA4, ICOS, TIGIT, IDO1*, and *TNFSF4*. These observations show that one dose of neoadjuvant pembrolizumab promoted enhanced inflammatory gene signatures in some patients without pTR, and raise the possibility that additional doses of neoadjuvant pembrolizumab, or addition of therapy targeting different immune checkpoints, could increase the proportion of patients with pTR. In preclinical models, targeting immune checkpoint inhibitor compensatory responses enabled bypass of single immune checkpoint inhibitor resistance.^28,29^

After neoadjuvant pembrolizumab, we observed tumor down-staging in 19% of patients. Also, the rate of high-risk pathology (50%) was slightly lower than historical (68%). These outcomes were unexpected, and must be confirmed in prospective controlled trials. The importance of these observations is that they could alter recommendations for adjuvant therapy. The best example of the potential to alter adjuvant therapy recommendations was patient 20, who presented with clinical stage IV (T2N2bM0) buccal SCC. After neoadjuvant pembrolizumab, the surgery specimen showed pathologic stage I (T1N0) disease. Extensive pTR was present, and only a 5mm residual focus of carcinoma. In the absence of adjuvant therapy, the patient remained free of disease 29 months after surgery. Neoadjuvant immunotherapy has the potential to reduce the intensity of adjuvant therapy in patients with resectable, locally advanced HNSCC.

Our trial has several limitations. The patient sample size was small, and the study design did not include a placebo-controlled arm. The proportion of patients with high-risk pathology was lower than expected, which resulted in fewer patients given adjuvant pembrolizumab. The standard of care adjuvant therapy varied. We cannot conclude the safety or benefit of adjuvant pembrolizumab given the limited sample size. Due to the limited size of pre-treatment biopsies, we were unable to perform multisector, targeted genome sequencing in baseline tumor samples. Several biomarkers correlated with achievement of major PTR to neoadjuvant pembrolizumab, but we were unable to address which biomarker was best suited to predict PTR. Relapse events through one-year after surgery were mature; however, longer follow-up is needed, because immunotherapy could delay the appearance of relapse events.

In conclusion, neoadjuvant pembrolizumab administered to patients with resectable locally advanced, HPV-unrelated HNSCC was safe, and resulted in pTR in 44% of patients and a one-year relapse rate lower than historical in patients with high-risk pathology. High-depth, multisector genome sequencing of surgical specimens documented tumor clonal loss after neoadjuvant pembrolizumab. Correlates of pTR with biomarkers of response and mechanisms of resistance to pembrolizumab were shown. Further studies of neoadjuvant pembrolizumab in patients with resectable locally advanced, HPV-unrelated HNSCC are warranted.

## Data Availability

Sequencing data have been deposited in dbGaP (phs001623).

## Contributors

All authors contributed to data collection, data interpretation and writing of the manuscript. KMC, AME, MG, JFP, DK, OLG, DA and RU designed the figures and did the data analysis. DA and RU wrote the clinical study design.

## Acknowledgements

We thank all patients and their families for their participation in this study. We recognize the support of the Alvin J. Siteman Cancer Center at Washington University School of Medicine and Barnes-Jewish Hospital in St. Louis, Missouri, the Clinical Trials Core, the Biostatistics Shared Resource, and the Center for Biomedical Informatics. The Siteman Cancer Center is supported in part by NCI Cancer Center Support Grant #P30 CA91842. MG is funded by the National Human Genome Research Institute (NIH NHGRI R00HG007940), OLG by the National Cancer Institute (NIH/NCI K22CA188163, NIH/NCI U01CA209936 and NIH/NCI U24CA237719) and a Cancer Research Foundation Young Investigator Award. RU is funded by the National Institute for Dental and Craniofacial Research (NIH/NIDCR R01DE024403, R01DE027736) and a V Foundation Translational Research Award. Clinical trial support was through a Merck Investigator Studies Program award to RU/DRA.

